# Polygenic risk scores for hypertensive disorders of pregnancy and later-life cardiovascular disease in women

**DOI:** 10.64898/2026.10.01.26364560

**Authors:** Gayathry Krishnamurthy, James A. Temple, Mustafa A. Ahmed, Jennie Hui, Peter J. Meikle, Corey Giles, Hoang T. Phan, Shaun P. Brennecke, Eric K. Moses, Phillip E. Melton

## Abstract

**Objectives:** To investigate whether genetic predisposition to hypertensive disorders of pregnancy (HDP), measured using polygenic risk scores (PRS), is associated with subsequent cardiovascular disease (CVD) outcomes in analyses stratified by HDP history.

**Study design and main outcome measures:** This population-based cohort included 1,284 women from the 1994-95 Busselton Health Study with genotype data, pregnancy history, cardiovascular risk factors, and linked hospitalisation and death records to 2022. Four HDP-related PRS were assessed: two for preeclampsia, one for gestational hypertension, and one for broad HDP phenotype. Outcomes were incident first CVD events classified into categories. Fine-Gray competing-risk models adjusted for age, cardiovascular risk factors, and genetic principal components estimated subdistribution hazard ratios (sHR) per standard deviation increase in PRS.

**Results:** Of 1,284 women, 309 reported a history of HDP and 651 experienced an incident CVD event during follow-up. In analyses stratified by HDP history, the largest estimates were observed for first cerebrovascular disease, although event numbers were small. The PGS003586 preeclampsia PRS showed the largest estimate (sHR 3.41, 95% confidence interval (CI) 1.54-7.58, p = 0.003), while PGS004593, another preeclampsia-related score, was also positively associated with cerebrovascular disease (sHR 1.86, 95% CI 1.05-3.30, p = 0.035). The gestational hypertension and broad HDP scores also showed positive estimates, but CI included 1.0, indicating inconclusive evidence.

**Conclusions:** Among women with a history of HDP, preeclampsia-related PRS showed preliminary evidence of an association with incident cerebrovascular disease. Given the small number of events, findings should be interpreted cautiously and validated in larger, multi-ancestry cohorts.

## 1. Introduction

Cardiovascular disease (CVD) is the leading cause of death among women worldwide, accounting for approximately 30% of female deaths [1]. Pregnancy may reveal underlying vascular and metabolic risk relevant to future maternal cardiovascular health [2]. Hypertensive disorders of pregnancy (HDP), including preeclampsia (PE) and gestational hypertension (GH), affect 5-10% of pregnancies globally [3,4] and are associated with increased later-life CVD risk [2,3,5-8].

Long-term CVD risk varies by HDP phenotype, severity, recurrence, and cardiovascular outcome [5-8]. PE is associated with an approximately four-fold higher risk of future heart failure and a two-fold higher risk of coronary heart disease, stroke, and cardiovascular death [8]. HDP and later-life CVD may share biological processes involving endothelial dysfunction, altered angiogenic signalling, inflammation, metabolic abnormalities, blood pressure dysregulation, and renal impairment [2,9-11]. HDP may therefore contribute to cardiovascular injury, reveal pre-existing maternal risk, or reflect both processes [2,9]. Shared genetic susceptibility has also been proposed [12-17], but its contribution to later-life CVD remains uncertain, particularly in longitudinal cohorts with pregnancy history, genotype data, cardiovascular risk factors, and linked clinical outcomes in the same women [12].

Polygenic risk scores (PRS) aggregate the weighted effects of multiple trait-associated variants into a measure of genetic predisposition [18]. PRS may complement conventional cardiovascular risk factors, although their incremental clinical value and transferability across ancestry groups require careful evaluation [18-21]. Recent PE- and GH-related PRS are associated with HDP across multiple cohorts [15,22], and emerging evidence suggests that genetic liability to HDP may also be associated with later-life CVD [23-25]. Despite this, it remains unclear whether HDP-related genetic risk is associated with specific future cardiovascular outcomes, particularly when other cardiovascular events may occur first and act as competing events [26-28].

Therefore, this study investigated whether polygenic risk for HDP identifies women at increased risk of specific later-life CVD outcomes. Using women from the Busselton Health Study (BHS), we evaluated PE-, GH-, and broad HDP-related PRS in analyses stratified by HDP history, while accounting for competing first cardiovascular events using Fine-Gray models.

## 2. Methods

### 2.1. Study population: Busselton Health Study (BHS)

Participants were included from the 1994-95 survey of the BHS, a long-running population-based cohort from Busselton, Western Australia, which commenced in 1966 and has repeatedly collected health data from predominantly European-ancestry individuals [29-32]. The 1994-95 survey included approximately 5,700 participants from earlier surveys and collected information on cardiometabolic risk factors, reproductive history, pregnancy-related conditions, and hormone use [29].

Women were eligible for the present analysis if they had genotype data, a history of pregnancy, available essential baseline covariates, consistent CVD information between questionnaires and linked records, and no prevalent CVD before baseline. The final analysis cohort included 1,284 women. Participant selection is summarised in Figure 1.

**Figure 1.**
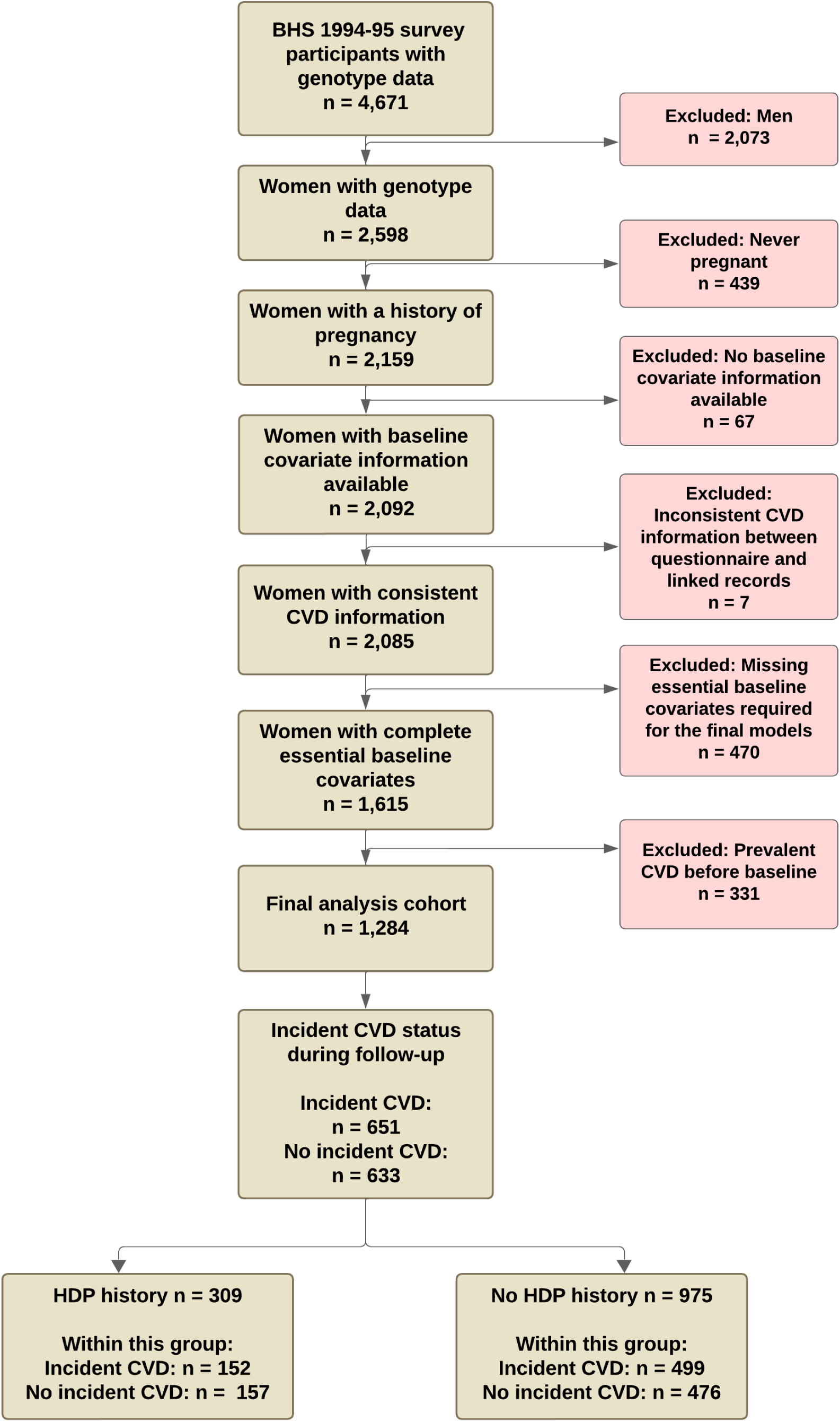
Participant selection flowchart. Flowchart showing the selection of women from the BHS 1994-95 survey into the final analysis cohort, with exclusions based on sex, pregnancy history, absence of baseline covariate information, inconsistent CVD information between questionnaire responses and linked health records, missing essential baseline covariates required for the final models, and prevalent CVD at baseline. Essential baseline covariates included age, SBP, WC, TC/HDL-C ratio, diabetes status, lipid-lowering medication use, and smoking status. The final cohort is also summarised by incident CVD status during follow-up and history of HDP. **Abbreviations:** BHS, Busselton Health Study; CVD, cardiovascular disease; HDL-C, high-density lipoprotein cholesterol; HDP, hypertensive disorders of pregnancy; SBP, systolic blood pressure; TC, total cholesterol; WC, waist circumference.

### 2.2. Definition of HDP and CVD

HDP history was based on participant self-report collected during the 1994-95 BHS survey. Women who reported hypertension, high blood pressure, or PE during any pregnancy were classified as having a history of HDP. Women who had been pregnant but did not report any of these conditions were classified as having no history of HDP. Further obstetric details, including the specific HDP subtype, timing of diagnosis, severity, treatment, and affected pregnancy, were not available.

CVD events were ascertained from linked hospitalisation and death records obtained from the Western Australian Department of Health Hospital Morbidity Data Collection and Death Registrations. Events recorded up to February 2023 were identified using International Classification of Diseases, Eighth, Ninth, and Tenth Revision codes.

The classification codes and broad category labels used to define each outcome are provided in Supplementary Table S1. The primary outcome was first incident CVD after the 1994-95 BHS baseline survey. First CVD outcomes were grouped as ischaemic heart disease, cerebrovascular disease, atrial fibrillation or flutter, heart failure, hypertensive diseases, peripheral vascular diseases, venous and lymphatic diseases, cardiac arrest, and ill-defined or unspecified circulatory diseases.

### 2.3. Baseline phenotypic variables

Baseline phenotypic variables included age, systolic blood pressure, waist circumference, total cholesterol to high-density lipoprotein cholesterol ratio, diabetes status, lipid-lowering medication use, and smoking status [30-32]. These covariates were selected based on cardiovascular relevance, alignment with previous risk prediction models, including the modified PREDICT New Zealand cardiovascular risk model, and model-fit assessment [32,33]. The covariate selection process, including comparison of candidate models using the Akaike information criterion, and details of phenotypic measurements are provided in Supplementary Methods S1.

### 2.4. Genotyping and imputation

Genotyping was performed using Illumina Human 610K Quad and Illumina 660W Quad arrays, with genotype quality control conducted as previously described [30,31]. Genomic coordinates were lifted to GRCh38/hg38 [36], and imputation was performed using the TOPMed Imputation Server following recommended protocols [37-39]. Post-imputation quality control and population structure assessment were conducted using standard procedures, with the first ten genetic principal components included in adjusted models [34,35]. Further details are provided in Supplementary Methods S2.

### 2.5. Polygenic risk score selection and calculation

Four HDP-related PRS were evaluated: two PE scores, PGS003586 [15] and PGS004593 [22]; one GH score, PGS003587 [15]; and one broad HDP score [24]. The two PE PRS and GH PRS were obtained from the Polygenic Score (PGS) Catalog [40,41] and calculated using pgsc_calc v2.2.0. The broad HDP PRS was obtained separately from its source publication and external repository and calculated using the published score weights [24,42]. Scores were standardised within the final analysis cohort, with effect estimates expressed per one standard deviation increase in PRS. Further details on score development, variant matching, calculation, and genetic ancestry assessment are provided in Supplementary Table S2 and Supplementary Methods S3.

### 2.6. Statistical analysis

#### 2.6.1. Descriptive statistics

Continuous variables were summarised as means and standard deviations and medians and interquartile ranges. Categorical variables, including first incident CVD outcomes, were summarised as frequencies and percentages. Between-group comparisons between women with and without a history of HDP used Welch’s two-sample t-test or the Wilcoxon rank-sum test for continuous variables and Pearson’s chi-square test or Fisher’s exact test for categorical variables, as appropriate. Descriptive comparison p-values were unadjusted.

#### 2.6.2. Follow-up definition and censoring

For time-to-event analyses, follow-up began at the 1994-95 BHS baseline survey and continued until the first recorded incident CVD event or the defined censoring time. Although linked hospitalisation and death-record data were available to February 2023, the administrative follow-up cut-off was the end of 2022, the last complete follow-up year. Participants without an incident CVD event were censored at death, where available, or 27 years after baseline.

#### 2.6.3. Competing-risk PRS analysis

Specific first CVD outcomes were examined to capture the initial cardiovascular manifestation and minimise the influence of preceding CVD events, subsequent treatment, and changes in clinical management. Fine-Gray competing-risk models were used because a different first CVD event precluded the outcome of interest from occurring first. By accounting for competing events rather than treating them as ordinary censoring, these models reduce potential overestimation of cumulative incidence and estimate associations with each first CVD outcome [26-28].

PRS were analysed as standardised continuous variables, with subdistribution hazard ratios (sHRs) and 95% confidence intervals (CIs) estimated per one standard deviation increase in each score. For each analysis, the CVD outcome of interest was treated as the event, other first CVD outcomes were treated as competing events, and participants without incident CVD were censored at their defined censoring time. Outcome categories with sparse event numbers among women with a history of HDP were excluded from the PRS association analyses.

Fully adjusted models included age, age^2^, systolic blood pressure, waist circumference, total cholesterol to high-density lipoprotein cholesterol ratio, diabetes status, lipid-lowering medication use, smoking status, and the first ten genetic PCs. Given the exploratory, hypothesis-generating nature of the study, no adjustment for multiple testing was applied. The PRS were selected a priori, and p-values <0.05 were interpreted as nominal evidence of association. Findings were considered alongside effect estimates, 95% CIs, event numbers, and consistency across scores and outcomes. Analyses were performed using R version 4.5.2 [43].

## 3. Results

### 3.1. Participant selection and baseline characteristics

Of 2,598 women with genotype data, 439 who had never been pregnant were excluded. Among the remaining 2,159 women, 67 were excluded because no baseline covariate information was available, seven because of inconsistent CVD information between questionnaires and linked records, 470 because of missing essential baseline covariates, and 331 because of prevalent CVD before baseline. The final analysis cohort comprised 1,284 women (Figure 1), including 309 with a history of HDP and 975 without HDP.

Baseline characteristics are presented in Table 1. Women with HDP were younger at baseline and had higher systolic blood pressure, waist circumference, and total cholesterol to high-density lipoprotein cholesterol ratio than women without HDP.

**Table 1.** Baseline characteristics by HDP history.

| Variables | Total cohort<br>(n = 1284) |  |  | HDP<br>(n = 309) |  |  | No HDP<br>(n = 975) |  |  | *P value |
| --- | --- | --- | --- | --- | --- | --- | --- | --- | --- | --- |
|  | Mean ± SD or<br>frequency (%) | Median | IQR | Mean ± SD<br>or frequency<br>(%) | Median | IQR | Mean ± SD or<br>frequency (%) | Median | IQR |  |
| Age (years) | 48.8 ± 14.5 | 45.0 | 22.9 | 46.6 ± 12.8 | 43.2 | 18.8 | 49.4 ± 14.9 | 45.6 | 23.9 | <0.001<sup>†</sup> |
| WC (cm) | 80.7 ± 11.4 | 78.3 | 14.0 | 83.0 ± 12.3 | 80.7 | 15.5 | 79.9 ± 11.0 | 77.5 | 13.6 | <0.001<sup>‡</sup> |
| TC/HDL-C ratio | 3.8 ± 1.2 | 3.6 | 1.5 | 4.0 ± 1.2 | 3.7 | 1.6 | 3.8 ± 1.2 | 3.5 | 1.5 | 0.009<sup>‡</sup> |
| SBP (mmHg) | 119.4 ± 17.5 | 116.7 | 22.7 | 122.3 ± 16.7 | 120.7 | 21.3 | 118.5 ± 17.7 | 115.3 | 22.0 | <0.001<sup>‡</sup> |
| Diabetes status,<br>n (%) | 24 (1.9%) |  |  | 8 (2.6%) |  |  | 16 (1.6%) |  |  | 0.284<sup>§</sup> |
| Lipid-lowering<br>medication use, n (%) | 21 (1.6%) |  |  | 4 (1.3%) |  |  | 17 (1.7%) |  |  | 0.588<sup>§</sup> |
| Smoking status, n (%) | 532 (41.4%) |  |  | 124 (40.1%) |  |  | 408 (41.8%) |  |  | 0.593<sup>§</sup> |

**Abbreviations:**
**WC**, waist circumference; **TC**, total cholesterol; **HDL-C**, high-density lipoprotein-cholesterol; **SBP**, systolic blood pressure; **HDP**, hypertensive disorders of pregnancy; **SD**, standard deviation; **IQR**, interquartile range.
Values are presented as mean ± standard deviation and median with interquartile range for continuous variables, and frequency (%) for categorical variables. IQR represents Q3-Q1. Welch's two-sample t-test or the Wilcoxon rank-sum test was used for continuous variables, as appropriate based on distributional characteristics. Pearson's chi-square test was used for categorical variables.
\*P values compare women with and without a history of HDP. <sup>†</sup>Welch's two-sample t-test; <sup>‡</sup>Wilcoxon rank-sum test; <sup>§</sup>Pearson's chi-square test. P values are unadjusted; p < 0.05 was considered nominally significant.

### 3.2. Distribution of first CVD outcomes

During follow-up, 651 women experienced an incident CVD event, including 152 with a history of HDP and 499 without HDP. The distribution of first CVD outcomes is presented in Table 2. Peripheral vascular diseases and cardiac arrest were excluded from subsequent PRS analyses because of sparse event numbers among women with HDP.

**Table 2:**
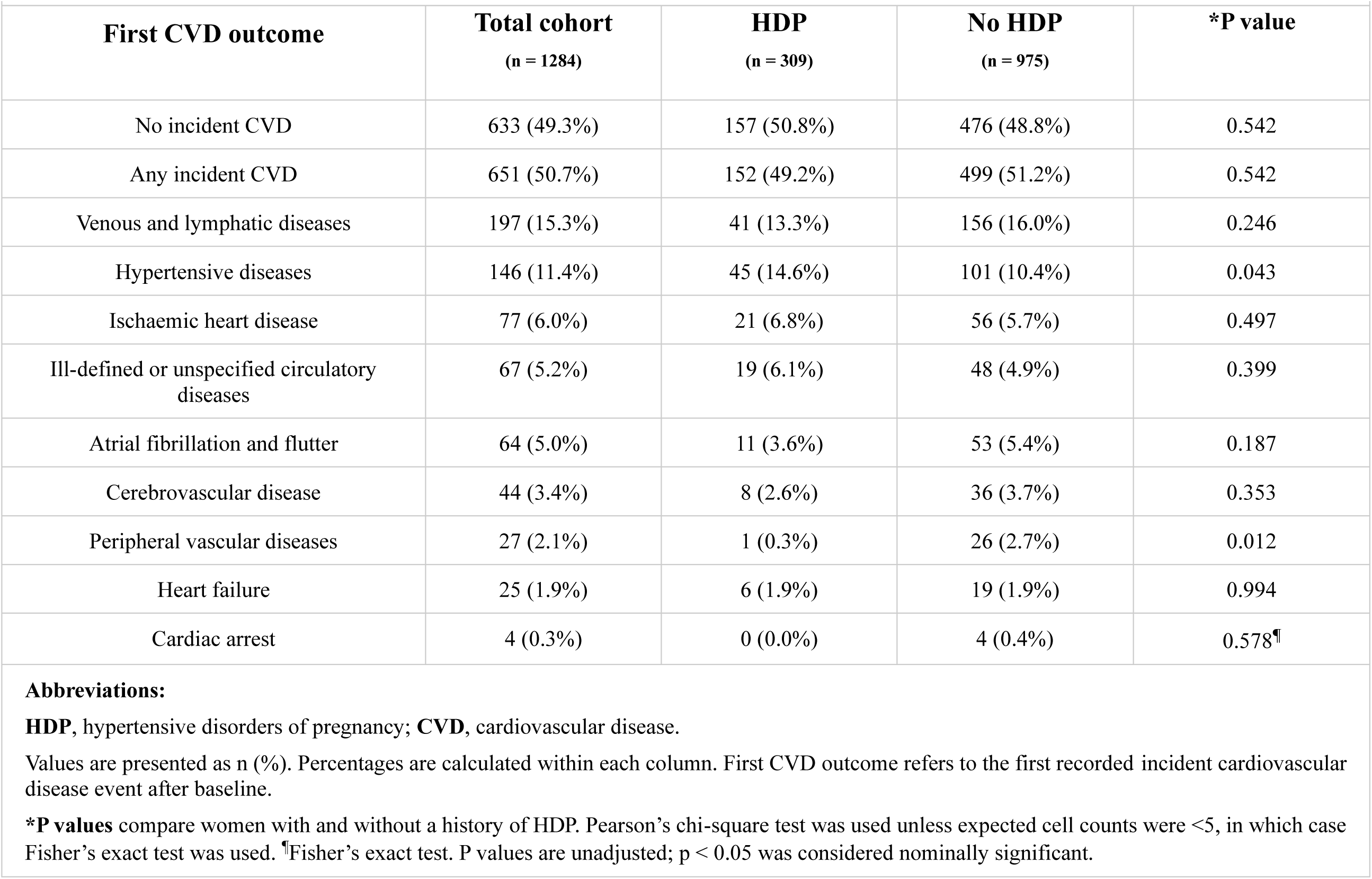
Distribution of first CVD outcomes by HDP history.

| Table 2: Distribution of first CVD outcomes by HDP history |  |  |  |  |
| --- | --- | --- | --- | --- |
| First CVD outcome | Total cohort<br>(n = 1284) | HDP<br>(n = 309) | No HDP<br>(n = 975) | *P value |
| No incident CVD | 633 (49.3%) | 157 (50.8%) | 476 (48.8%) | 0.542 |
| Any incident CVD | 651 (50.7%) | 152 (49.2%) | 499 (51.2%) | 0.542 |
| Venous and lymphatic diseases | 197 (15.3%) | 41 (13.3%) | 156 (16.0%) | 0.246 |
| Hypertensive diseases | 146 (11.4%) | 45 (14.6%) | 101 (10.4%) | 0.043 |
| Ischaemic heart disease | 77 (6.0%) | 21 (6.8%) | 56 (5.7%) | 0.497 |
| Ill-defined or unspecified circulatory diseases | 67 (5.2%) | 19 (6.1%) | 48 (4.9%) | 0.399 |
| Atrial fibrillation and flutter | 64 (5.0%) | 11 (3.6%) | 53 (5.4%) | 0.187 |
| Cerebrovascular disease | 44 (3.4%) | 8 (2.6%) | 36 (3.7%) | 0.353 |
| Peripheral vascular diseases | 27 (2.1%) | 1 (0.3%) | 26 (2.7%) | 0.012 |
| Heart failure | 25 (1.9%) | 6 (1.9%) | 19 (1.9%) | 0.994 |
| Cardiac arrest | 4 (0.3%) | 0 (0.0%) | 4 (0.4%) | 0.578<sup>†</sup> |
| **Abbreviations:**<br>**HDP**, hypertensive disorders of pregnancy; **CVD**, cardiovascular disease.<br>Values are presented as n (%). Percentages are calculated within each column. First CVD outcome refers to the first recorded incident cardiovascular disease event after baseline.<br>**\*P values** compare women with and without a history of HDP. Pearson's chi-square test was used unless expected cell counts were <5, in which case Fisher's exact test was used. <sup>†</sup>Fisher's exact test. P values are unadjusted; p < 0.05 was considered nominally significant. |  |  |  |  |

### 3.3. Associations between HDP-related PRS and first CVD outcomes

Associations between HDP-related PRS and first CVD outcomes are presented in Figures 2 and 3. Among women with a history of HDP, the strongest associations were observed for first cerebrovascular disease. Although only eight cerebrovascular events were observed in this group, the PGS003586 PE PRS was associated with an increased risk of first cerebrovascular disease (sHR 3.41, 95% CI 1.54-7.58, p = 0.003). A second PE PRS, PGS004593, also showed a positive association with first cerebrovascular disease (sHR 1.86, 95% CI 1.05-3.30, p = 0.035).

**Figure 2.**
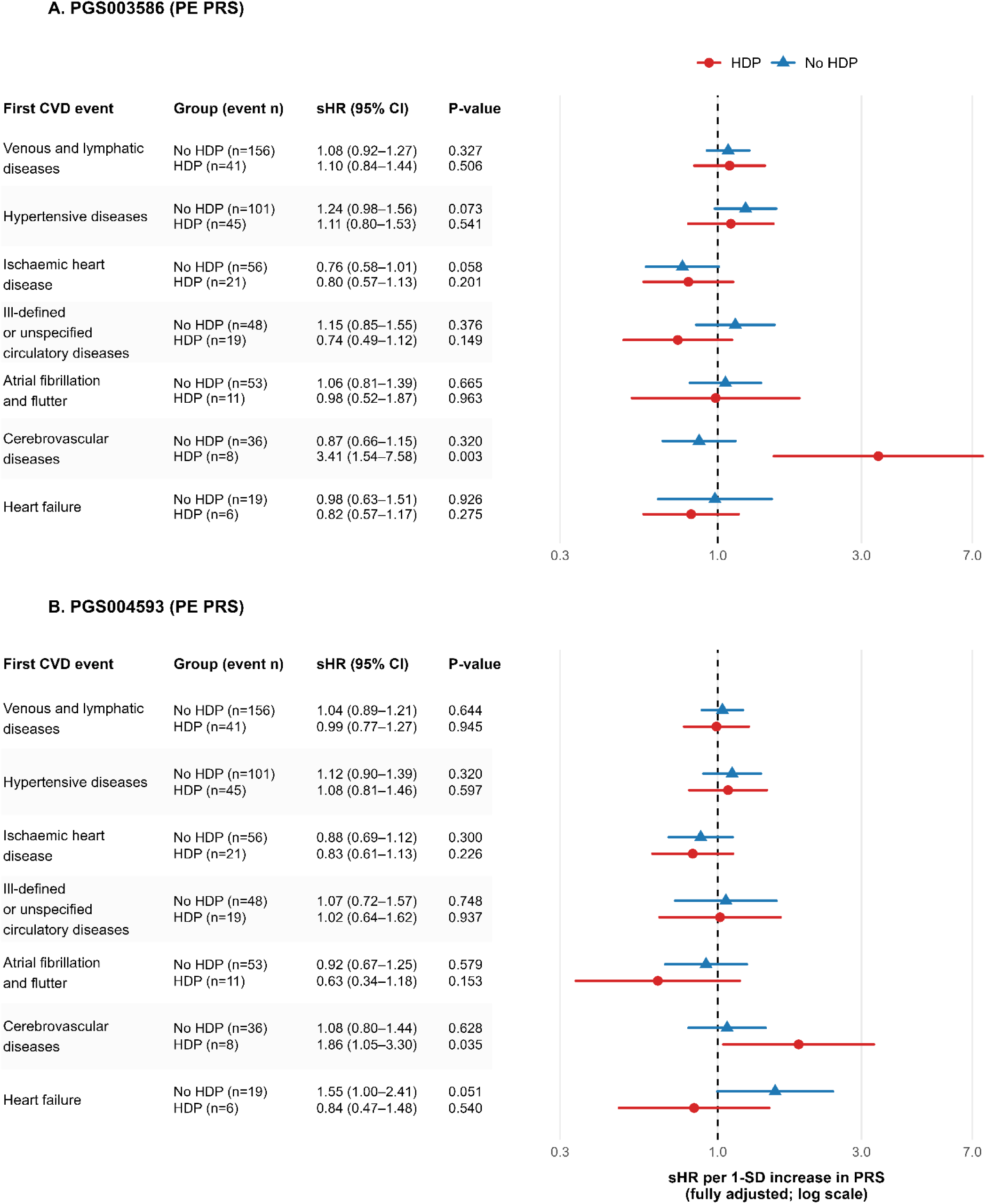
Associations of PE PRS with later-life first CVD outcomes by HDP history. Forest plots show associations between each PE PRS and first CVD outcomes, stratified by HDP history. Panel A shows PGS003586 and Panel B shows PGS004593. Estimates are sHRs per 1-SD increase in PRS from fully adjusted Fine-Gray competing-risk models. Models were adjusted for age, age^2^, SBP, WC, TC/HDL-C ratio, diabetes status, lipid-lowering medication use, smoking status and the first 10 genetic PCs. Points represent sHRs and horizontal lines represent 95% CIs. Group event numbers are shown in the table. P-values represent the within-stratum association between each PRS and first CVD outcome among women with and without HDP history; nominal significance was defined as p < 0.05. **Abbreviations:** CI, confidence interval; CVD, cardiovascular disease; HDL-C, high-density lipoprotein cholesterol; HDP, hypertensive disorders of pregnancy; PCs, principal components; PE, preeclampsia; PRS, polygenic risk score; SBP, systolic blood pressure; SD, standard deviation; sHR, subdistribution hazard ratio; TC, total cholesterol; WC, waist circumference.

**Figure 3.**
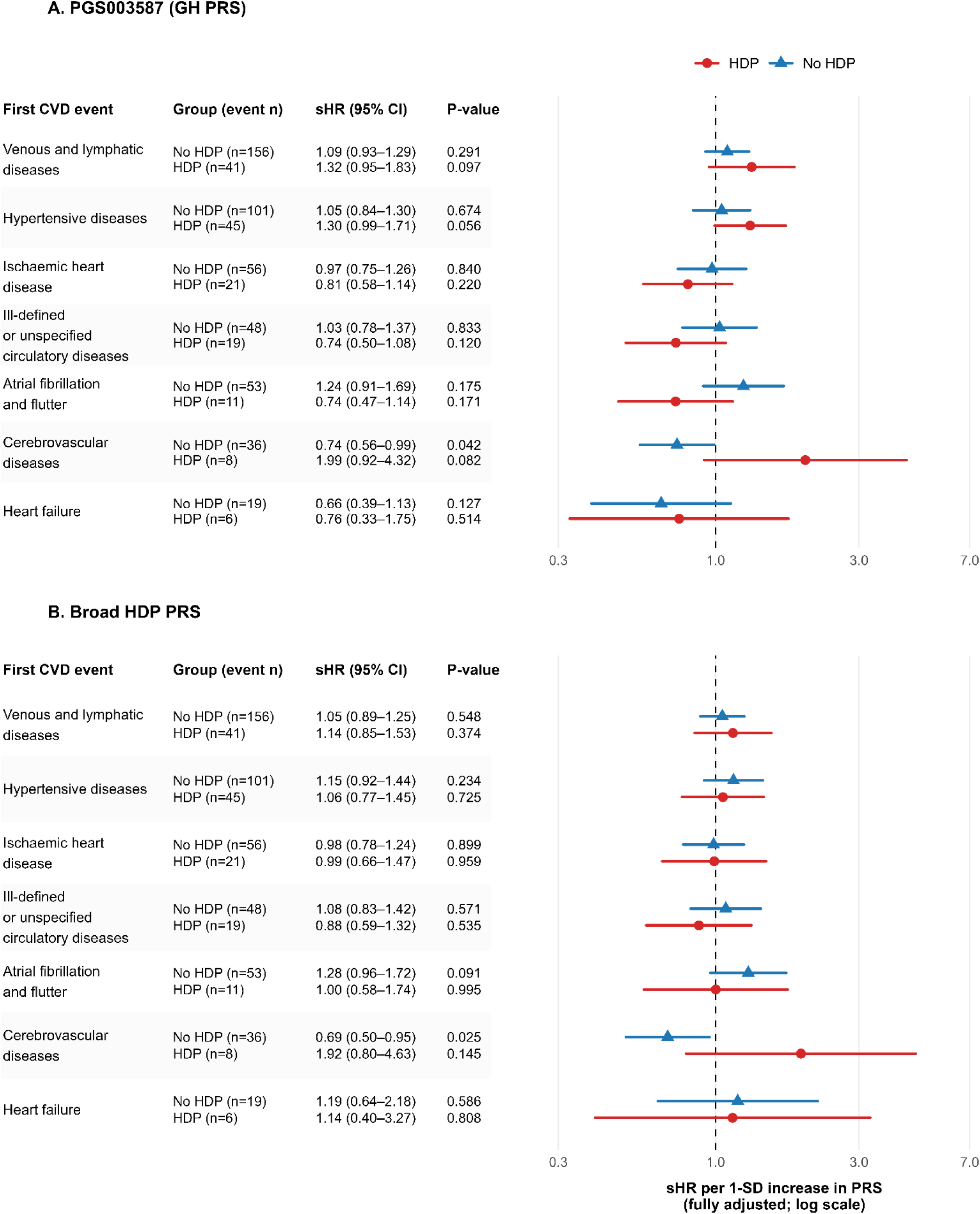
Associations of GH PRS and broad HDP PRS with later-life first CVD outcomes by HDP history. Forest plots show associations of the GH PRS and broad HDP PRS with first CVD outcomes, stratified by HDP history. Panel A shows PGS003587 and Panel B shows the broad HDP PRS. Estimates are sHRs per 1-SD increase in PRS from fully adjusted Fine-Gray competing-risk models. Models were adjusted for age, age^2^, SBP, WC, TC/HDL-C ratio, diabetes status, lipid-lowering medication use, smoking status, and the first 10 genetic PCs. Points represent sHRs and horizontal lines represent 95% CIs. Group event numbers are shown in the table. P-values represent the within-stratum association between each PRS and first CVD outcome among women with and without HDP history; nominal significance was defined as p < 0.05. The broad HDP PRS was obtained from an external source repository and is not a PGS Catalog score. **Abbreviations:** CI, confidence interval; CVD, cardiovascular disease; GH, gestational hypertension; HDL-C, high-density lipoprotein cholesterol; HDP, hypertensive disorders of pregnancy; PCs, principal components; PRS, polygenic risk score; SBP, systolic blood pressure; SD, standard deviation; sHR, subdistribution hazard ratio; TC, total cholesterol; WC, waist circumference.

The GH PRS (PGS003587) and the broad HDP PRS showed similar positive estimates for first cerebrovascular disease, although their CI included the null (sHR 1.99, 95% CI 0.92-4.32, p = 0.082; and sHR 1.92, 95% CI 0.80-4.63, p = 0.145, respectively). Associations with other first CVD outcomes were generally weaker and less consistent across scores, with CI frequently including the null.

## 4. Discussion

In this population-based cohort of women with long-term follow-up, genetic susceptibility to HDP, particularly PE, was the most strongly associated with subsequent cerebrovascular disease. Two independent PE PRS (PGS003586, PGS004593), demonstrated increased risk with incident cerebrovascular disease among women with a history of HDP, while associations with other cardiovascular outcomes were generally weaker. These findings suggest that the genetic architecture underlying HDP may have a stronger relationship with later cerebrovascular risk than with other CVD outcomes, highlighting potential heterogeneity in the pathways linking pregnancy complications and future CVD risk.

The stronger associations observed for cerebrovascular disease may reflect biological pathways shared between HDP and cerebrovascular pathology. PE and cerebrovascular disease are both characterised by blood pressure dysregulation, endothelial dysfunction, altered angiogenic signalling, vascular inflammation, metabolic dysfunction, and renal impairment [2,9-11]. These shared mechanisms have been implicated in both, the development of HDP and the long-term risk of stroke and other cerebrovascular outcomes [44]. Brohan et al. reported increased long-term risks of any, ischaemic, and haemorrhagic stroke after HDP [5], while meta-analytic evidence showed an approximately two-fold higher risk of stroke after PE [8]. Examining first cardiovascular presentations may also have helped identify this pattern by preserving event ordering and reducing the influence of preceding events and subsequent clinical management.

A previous study by Lee et al. reported that high HDP genetic risk was associated with atherosclerotic CVD, coronary artery disease, myocardial infarction, and peripheral artery disease, but not ischaemic stroke [23]. In contrast, the strongest associations in the present study were observed for cerebrovascular disease among women with a history of HDP. These differences may reflect variation in outcome definitions, study populations, and analytical approaches. Whereas Lee et al. examined specific cardiovascular endpoints, we focused on first cardiovascular presentations within a competing-risk framework and evaluated both HDP-related and PE-specific PRS. It is therefore possible that considering event ordering and competing CVD outcomes may reveal disease-specific associations that are less apparent in conventional time-to-event analyses.

These findings support further investigation of HDP-related PRS as markers of shared genetic pathways between pregnancy hypertension and later CVD. The stronger associations observed for cerebrovascular disease raise the possibility that HDP-related genetic risk may not be distributed uniformly across cardiovascular outcomes but instead may preferentially influence specific disease pathways. Recent evidence suggests that high genetic risk for PE and CVD-related traits may further increase long-term CVD risk among women with PE or any HDP [25]. However, larger studies are required to determine whether these HDP-related PRS provide clinically meaningful information beyond established cardiovascular risk factors and pregnancy history. At present, these PRS are best viewed as tools for investigating shared biological mechanisms and refining risk prediction, rather than as stand-alone clinical predictors [18,19,23-25].

The present study has several strengths. The BHS provided a well-characterised population-based cohort with reproductive history, genotype data, baseline cardiovascular risk factors, and long-term linked hospitalisation and death-record follow-up [29-32]. Exclusion of women with prevalent CVD ensured that analyses focused on incident CVD outcomes. In addition, the competing-risk framework enabled evaluation of first CVD events while accounting for alternative CVD outcomes that could preclude their occurrence [26-28]. Analyses were adjusted for established cardiovascular risk factors and population structure, and all PRS demonstrated high compatibility with the imputed genetic dataset supporting the robustness of the genetic analyses.

Several limitations should be noted. HDP history was self-reported, which may have introduced recall error and exposure misclassification. In addition, the available data did not allow detailed classification by HDP subtype, timing, severity, or recurrence. As HDP exposure in the BHS was defined as an overall history of pregnancy hypertension, PE and GH could not be examined as separate clinical exposure groups. However, HDP history was recorded before follow-up for incident CVD, and women with prevalent CVD were excluded, reducing the potential for differential misclassification related to subsequent CVD outcomes. Any remaining misclassification of HDP status was likely to be largely non-differential with respect to later incident CVD and may have attenuated the observed associations.

The small number of cerebrovascular events relative to model covariates may have resulted in imprecise and potentially inflated effect estimates. Accordingly, the large effects observed for PGS003586 should be interpreted as an exploratory signal rather than a stable estimate of clinical risk. In addition, multiple PRS-outcome associations were examined without formal adjustment for multiple testing, and the reported p-values therefore represent nominal evidence of association. Nevertheless, the consistent direction of effect across all four HDP-related PRS for cerebrovascular disease, together with the use of externally developed scores and a competing-risk framework, supports further evaluation in independent cohorts.

The cohort was predominantly European ancestry, which improves comparability with the largely European-derived PRS included in this study but may limit generalisability to other populations [15,18,19,22,24]. In addition, CVD outcomes were grouped into broad categories based on hospitalisation and death records, potentially encompassing clinically heterogeneous conditions. Finally, the use of complete-case analysis may have introduced selection bias if participants because of missing data differed systematically from those included in the final analyses.

In conclusion, PE-related PRS were associated with an increased risk of first cerebrovascular disease among women with a history of HDP, whereas associations with other CVD outcomes were generally weaker. These findings support the possibility that genetic liability to HDP may contribute differentially across CVD outcomes and are consistent with existing evidence linking PE to later cerebrovascular risk. However, given the small number of cerebrovascular events, the findings should be considered exploratory and require validation in larger, more diverse cohorts with well-characterised pregnancy histories. Future studies should also determine whether HDP-related PRS provide clinically meaningful information beyond established cardiovascular risk factors and pregnancy history.

## Abbreviations

CVD: cardiovascular disease
CI: confidence interval
GH: gestational hypertension
HDP: hypertensive disorders of pregnancy
PCs: principal components
PE: preeclampsia
PGS: polygenic score
PRS: polygenic risk score
sHR: subdistribution hazard ratio

## Code Availability

Code supporting the polygenic risk score calculation and Fine-Gray competing-risk analyses is available on GitHub: https://github.com/UTASCVD-genomics/HDP-CVD-PRS.

## Data Availability

The data used in this study are subject to ethical, governance and data-custodian restrictions and are not publicly available. Access to Busselton Health Study and linked health data may be obtained through the relevant data custodians subject to appropriate approvals.

## Ethics Approval

The Human Research Ethics Committee (HREC) of the University of Tasmania granted ethics approval on September 27, 2021, for the Australian Genomics of Cardiovascular Disease Risk Following Preeclampsia Study (Project ID: 24058). Informed consent was obtained from all participants.

## Author contribution

**GK:** Conceptualisation, Methodology, Formal analysis, Investigation, Data curation, Visualisation, Writing - original draft. **JAT:** Methodology. **MAA:** Methodology, Data curation. **JH:** Resources. **PJM, CG:** Resources, Data curation. **HTP:** Supervision, Writing - review and editing. **SPB:** Methodology, Writing - review and editing. **EKM:** Supervision, Funding acquisition, Writing - review and editing. **PEM:** Conceptualisation, Methodology, Supervision, Project administration, Funding acquisition, Writing - review and editing.

## Declaration of competing interest

The authors declare that they have no competing financial interests or personal relationships that could have influenced the work presented in this article.

## Funding

All financial support for this study was provided by the National Health and Medical Research Council (NHMRC), Australia (Grants APP2001203 and APP2028555), the Tasmania Graduate Research Scholarship (TGRS) from the University of Tasmania, Hobart, Australia, and the Menzies Institute for Medical Research Top-up Scholarship.

## Acknowledgements

We gratefully acknowledge the NHMRC, TGRS, and the Menzies Institute for Medical Research for their funding support. We also thank the Menzies Institute for Medical Research, University of Tasmania, for providing access to research facilities and resources.

We extend our gratitude to the Busselton Health Study and the 1994-95 survey participants for their invaluable contributions. We acknowledge the Pawsey Supercomputing Research Centre for providing computational resources that enabled the analyses, the Western Australia Health Linked Data for providing CVD outcome data, and PathWest Laboratory Medicine, WA, for genotyping services. Finally, we thank all staff and collaborators involved in data collection, management, and support throughout this study.

## Notes

### Competing Interest Statement

The authors have declared no competing interest.

## References

[1] World Heart Federation, Women & CVD. https://world-heart-federation.org/heart-health/women-cvd/ (accessed 29 July 2026).

[2] J.W. Rich-Edwards, A. Fraser, D.A. Lawlor, J.M. Catov, Pregnancy characteristics and women’s future cardiovascular health: an underused opportunity to improve women’s health?, Epidemiol. Rev. 36 (2014) 57–70. 10.1093/epirev/mxt006.

[3] A.M. Khedagi, N.A. Bello, Hypertensive disorders of pregnancy, Cardiol. Clin. 39 (2021) 77–90. 10.1016/j.ccl.2020.09.005.

[4] L.A. Magee, M.A. Brown, D.R. Hall, S. Gupte, A. Hennessy, S.A. Karumanchi, et al., The 2021 International Society for the Study of Hypertension in Pregnancy classification, diagnosis & management recommendations for international practice, Pregnancy Hypertens. 27 (2022) 148–169. 10.1016/j.preghy.2021.09.008.

[5] M.P. Brohan, F.P. Daly, L. Kelly, F.P. McCarthy, A.S. Khashan, K. Kublickiene, et al., Hypertensive disorders of pregnancy and long-term risk of maternal stroke-a systematic review and meta-analysis, Am. J. Obstet. Gynecol. 229 (2023) 248–268.e7. 10.1016/j.ajog.2023.03.034.

[6] L. Brouwers, A.J. van der Meiden-van Roest, C. Savelkoul, T.E. Vogelvang, A.T. Lely, A. Franx, et al., Recurrence of pre-eclampsia and the risk of future hypertension and cardiovascular disease: a systematic review and meta-analysis, BJOG 125 (2018) 1642–1654. 10.1111/1471-0528.15394.

[7] A. Dall’Asta, F. D’Antonio, G. Saccone, D. Buca, E. Mastantuoni, M. Liberati, et al., Cardiovascular events following pregnancy complicated by pre-eclampsia with emphasis on comparison between early- and late-onset forms: systematic review and meta-analysis, Ultrasound Obstet. Gynecol. 57 (2021) 698–709. 10.1002/uog.22107.

[8] P. Wu, R. Haththotuwa, C.S. Kwok, A. Babu, R.A. Kotronias, C. Rushton, et al., Preeclampsia and future cardiovascular health: a systematic review and meta-analysis, Circ. Cardiovasc. Qual. Outcomes 10 (2017) e003497. 10.1161/CIRCOUTCOMES.116.003497.

[9] S. Rana, E. Lemoine, J.P. Granger, S.A. Karumanchi, Preeclampsia: pathophysiology, challenges, and perspectives, Circ. Res. 124 (2019) 1094-1112. 10.1161/CIRCRESAHA.118.313276.

[10] R.J. Levine, S.E. Maynard, C. Qian, K.H. Lim, L.J. England, K.F. Yu, et al., Circulating angiogenic factors and the risk of preeclampsia, N. Engl. J. Med. 350 (2004) 672–683. 10.1056/NEJMoa031884.

[11] S.E. Maynard, J.Y. Min, J. Merchan, K.H. Lim, J. Li, S. Mondal, et al., Excess placental soluble fms-like tyrosine kinase 1 (sFlt1) may contribute to endothelial dysfunction, hypertension, and proteinuria in preeclampsia, J. Clin. Invest. 111 (2003) 649–658. 10.1172/JCI17189.

[12] G. Krishnamurthy, P.T. Nguyen, B.N. Tran, H.T. Phan, S.P. Brennecke, E.K. Moses, et al., Genomic variation associated with cardiovascular disease progression following preeclampsia: a systematic review, Front. Epidemiol. 3 (2023) 1221222. 10.3389/fepid.2023.1221222.

[13] M.P. Johnson, S.P. Brennecke, C.E. East, T.D. Dyer, L.T. Roten, J.M. Proffitt, et al., Genetic dissection of the pre-eclampsia susceptibility locus on chromosome 2q22 reveals shared novel risk factors for cardiovascular disease, Mol. Hum. Reprod. 19 (2013) 423–437. 10.1093/molehr/gat011.

[14] J.S. Tyrmi, T. Kaartokallio, A.I. Lokki, T. Jääskeläinen, E. Kortelainen, S. Ruotsalainen, et al., Genetic risk factors associated with preeclampsia and hypertensive disorders of pregnancy, JAMA Cardiol. 8 (2023) 674–683. 10.1001/jamacardio.2023.1312.

[15] M.C. Honigberg, B. Truong, R.R. Khan, B. Xiao, L. Bhatta, H.M.T. Vy, et al., Polygenic prediction of preeclampsia and gestational hypertension, Nat. Med. 29 (2023) 1540–1549. 10.1038/s41591-023-02374-9.

[16] R. McGinnis, V. Steinthorsdottir, N.O. Williams, G. Thorleifsson, S. Shooter, S. Hjartardottir, et al., Variants in the fetal genome near FLT1 are associated with risk of preeclampsia, Nat. Genet. 49 (2017) 1255–1260. 10.1038/ng.3895.

[17] V. Steinthorsdottir, R. McGinnis, N.O. Williams, L. Stefansdottir, G. Thorleifsson, S. Shooter, et al., Genetic predisposition to hypertension is associated with preeclampsia in European and Central Asian women, Nat. Commun. 11 (2020) 5976. 10.1038/s41467-020-19733-6.

[18] S.W. Choi, T.S.H. Mak, P.F. O’Reilly, Tutorial: a guide to performing polygenic risk score analyses, Nat. Protoc. 15 (2020) 2759–2772. 10.1038/s41596-020-0353-1.

[19] J.W. O’Sullivan, S. Raghavan, C. Marquez-Luna, J.A. Luzum, S.M. Damrauer, E.A. Ashley, et al., Polygenic risk scores for cardiovascular disease: a scientific statement from the American Heart Association, Circulation 146 (2022) e93–e118. 10.1161/CIR.0000000000001077.

[20] N. Mars, J.T. Koskela, P. Ripatti, T.T.J. Kiiskinen, A.S. Havulinna, J.V. Lindbohm, et al., Polygenic and clinical risk scores and their impact on age at onset and prediction of cardiometabolic diseases and common cancers, Nat. Med. 26 (2020) 549–557. 10.1038/s41591-020-0800-0.

[21] M. Inouye, G. Abraham, C.P. Nelson, A.M. Wood, M.J. Sweeting, F. Dudbridge, et al., Genomic risk prediction of coronary artery disease in 480,000 adults: implications for primary prevention, J. Am. Coll. Cardiol. 72 (2018) 1883–1893. 10.1016/j.jacc.2018.07.079.

[22] J. Nurkkala, A. Kauko, FinnGen, H. Laivuori, T. Saarela, J.S. Tyrmi, et al., Associations of polygenic risk scores for preeclampsia and blood pressure with hypertensive disorders of pregnancy, J. Hypertens. 41 (2023) 380–387. 10.1097/HJH.0000000000003336.

[23] S.M. Lee, M. Shivakumar, B. Xiao, S.H. Jung, Y. Nam, J.S. Yun, et al., Genome-wide polygenic risk scores for hypertensive disease during pregnancy can also predict the risk for long-term cardiovascular disease, Am. J. Obstet. Gynecol. 229 (2023) 298.e1-298.e19. 10.1016/j.ajog.2023.03.013.

[24] S.H. Jung, H. Kim, Y.M. Jung, M. Shivakumar, B. Xiao, J. Kim, et al., Healthy lifestyle reduces cardiovascular risk in women with genetic predisposition to hypertensive disorders of pregnancy, Nat. Commun. 16 (2025) 1463. 10.1038/s41467-025-56107-2.

[25] A. Kivioja, J. Tyrmi, E. Toivonen, H. Huhtala, FinnGen, T. Jääskeläinen, et al., Long-term cardiovascular risk in women with hypertensive disorders of pregnancy: insights from polygenic risk scores, Acta Obstet. Gynecol. Scand. 104 (2025) 1907–1917. 10.1111/aogs.70021.

[26] J.P. Fine, R.J. Gray, A proportional hazards model for the subdistribution of a competing risk, J. Am. Stat. Assoc. 94 (1999) 496–509. 10.1080/01621459.1999.10474144.

[27] P.C. Austin, J.P. Fine, Practical recommendations for reporting Fine-Gray model analyses for competing risk data, Stat. Med. 36 (2017) 4391–4400. 10.1002/sim.7501.

[28] P.C. Austin, D.S. Lee, J.P. Fine, Introduction to the analysis of survival data in the presence of competing risks, Circulation 133 (2016) 601–609. 10.1161/CIRCULATIONAHA.115.017719.

[29] Busselton Population Medical Research Institute (BPMRI), Busselton Health Study. https://bpmri.org.au/ (accessed 29 July 2026).

[30] G. Cadby, P.E. Melton, N.S. McCarthy, M. Almeida, S. Williams-Blangero, J.E. Curran, et al., Pleiotropy of cardiometabolic syndrome with obesity-related anthropometric traits determined using empirically derived kinships from the Busselton Health Study, Hum. Genet. 137 (2018) 45–53. 10.1007/s00439-017-1856-x.

[31] G. Cadby, C. Giles, P.E. Melton, K. Huynh, N.A. Mellett, T. Duong, et al., Comprehensive genetic analysis of the human lipidome identifies loci associated with lipid homeostasis with links to coronary artery disease, Nat. Commun. 13 (2022) 3124. 10.1038/s41467-022-30875-7.

[32] M.W. Knuiman, J. Hung, M.L. Divitini, T.M.E. Davis, J.P. Beilby, Utility of the metabolic syndrome and its components in the prediction of incident cardiovascular disease: a prospective cohort study, Eur. J. Cardiovasc. Prev. Rehabil. 16 (2009) 235–241. 10.1097/HJR.0b013e32832955fc.

[33] R. Pylypchuk, S. Wells, A. Kerr, K. Poppe, T. Riddell, M. Harwood, et al., Cardiovascular disease risk prediction equations in 400 000 primary care patients in New Zealand: a derivation and validation study, Lancet 391 (2018) 1897–1907. 10.1016/S0140-6736(18)30664-0.

[34] S. Purcell, B. Neale, K. Todd-Brown, L. Thomas, M.A.R. Ferreira, D. Bender, et al., PLINK: a tool set for whole-genome association and population-based linkage analyses, Am. J. Hum. Genet. 81 (2007) 559–575. 10.1086/519795.

[35] C.C. Chang, C.C. Chow, L.C.A.M. Tellier, S. Vattikuti, S.M. Purcell, J.J. Lee, Second-generation PLINK: rising to the challenge of larger and richer datasets, Gigascience 4 (2015) 7. 10.1186/s13742-015-0047-8.

[36] A.S. Hinrichs, D. Karolchik, R. Baertsch, G.P. Barber, G. Bejerano, H. Clawson, et al., The UCSC Genome Browser Database: update 2006, Nucleic Acids Res. 34 (2006) D590–D598. 10.1093/nar/gkj144.

[37] D. Taliun, D.N. Harris, M.D. Kessler, J. Carlson, Z.A. Szpiech, R. Torres, et al., Sequencing of 53,831 diverse genomes from the NHLBI TOPMed Program, Nature 590 (2021) 290–299. 10.1038/s41586-021-03205-y.

[38] P.R. Loh, P. Danecek, P.F. Palamara, C. Fuchsberger, Y.A. Reshef, H.K. Finucane, et al., Reference-based phasing using the Haplotype Reference Consortium panel, Nat. Genet. 48 (2016) 1443–1448. 10.1038/ng.3679.

[39] S. Das, L. Forer, S. Schönherr, C. Sidore, A.E. Locke, A. Kwong, et al., Next-generation genotype imputation service and methods, Nat. Genet. 48 (2016) 1284–1287. 10.1038/ng.3656.

[40] S.A. Lambert, L. Gil, S. Jupp, S.C. Ritchie, Y. Xu, A. Buniello, et al., The Polygenic Score Catalog as an open database for reproducibility and systematic evaluation, Nat. Genet. 53 (2021) 420–425. 10.1038/s41588-021-00783-5.

[41] S.A. Lambert, B. Wingfield, J.T. Gibson, L. Gil, S. Ramachandran, F. Yvon, et al., Enhancing the Polygenic Score Catalog with tools for score calculation and ancestry normalization, Nat. Genet. 56 (2024) 1989–1994. 10.1038/s41588-024-01937-x.

[42] Dokyoon Kim Lab, PRS model for hypertensive disorders of pregnancy: hdp-prs-finngen-r8, Zenodo. 10.5281/zenodo.14211027 (accessed 29 July 2026).

[43] R Core Team, R: A language and environment for statistical computing, R Foundation for Statistical Computing, Vienna, Austria, 2025. https://www.R-project.org/.

[44] E.C. Miller, Preeclampsia and cerebrovascular disease: the maternal brain at risk, Hypertension 74 (2019) 5–13. 10.1161/HYPERTENSIONAHA.118.11513.

