## Supplementary Materials for "Polygenic risk scores for hypertensive disorders of pregnancy and later-life cardiovascular disease in women"

**Appendix A. Supplementary Materials**

| **Supplementary Table S1. ICD categories used to define first incident CVD outcome group** | | | | |
| --- | --- | --- | --- | --- |
| **First CVD outcome group** | **Broad ICD category labels grouped into this outcome** | **ICD-10 codes** | **ICD-9 codes** | **ICD-8 codes** |
| Hypertensive diseases | Hypertensive diseases | I10-I15 | 401-405 | 401-404 |
| Ischaemic heart disease | Ischaemic heart diseases | I20-I25 | 410-414 | 410-414 |
| Heart failure | Heart failure; congestive heart failure; left ventricular failure | I50 | 428 | 427.0, 427.1 |
| Ill-defined/unspecified circulatory diseases | Complications and ill-defined descriptions of heart disease; ill-defined descriptions and complications of heart disease; other and unspecified disorders of the circulatory system | I51, I95-I99 | 429 | 429 |
| Cerebrovascular disease | Cerebrovascular disease | I60-I69 | 430-438 | 430-438 |
| Peripheral vascular diseases | Diseases of arteries, arterioles and capillaries | I70-I79 | 440-449 | 440-444, 448, 456 |
| Venous and lymphatic diseases | Diseases of veins and lymphatics, and other diseases of circulatory system; diseases of veins, lymphatic vessels and lymph nodes, not elsewhere classified | I80-I89 | 451-459* | 451-457* |
| Atrial fibrillation and flutter | Atrial fibrillation and flutter | I48 | 427.3 | 427.4, 427.9 |
| Cardiac arrest | Cardiac arrest | I46 | 427.5 | 427.2 |

**Note:** First CVD outcome groups were defined using the ICD code and broad ICD category label corresponding to each participant’s first recorded incident CVD event after baseline. Outcome groups reflect the ICD category labels available in the linked hospitalisation and death-record data. *Older ICD-8 and ICD-9 category headings group diseases of veins and lymphatics together with some other circulatory disorders; this historical broad category was retained under the venous and lymphatic diseases outcome group. The category “ill-defined/unspecified circulatory diseases” included records labelled as complications or ill-defined descriptions of heart disease, as well as records labelled as other or unspecified disorders of the circulatory system. Each first CVD event was assigned to one mutually exclusive outcome group. **CVD**, cardiovascular disease; **ICD**, International Classification of Diseases.

**Supplementary Methods**

**Supplementary Methods S1. Phenotypic measurements and covariate selection**

Phenotypic data from the 1994-95 Busselton Health Study (BHS) survey included age, anthropometric measures, lipid measures, blood pressure measurements, glucose metabolism traits, medical history, medication use, and lifestyle variables. Age was recorded in years. Anthropometric measures included height (cm), weight (kg), body mass index (BMI; kg/m²), waist circumference, hip circumference, and waist-to-hip ratio. BMI was calculated from measured height and weight. Waist circumference was measured twice and averaged for analysis.

Lipid measures included total cholesterol (TC; mmol/L), low-density lipoprotein cholesterol (LDL-C; mmol/L), high-density lipoprotein cholesterol (HDL-C; mmol/L), TC/HDL-C ratio, and triglycerides (mmol/L). Fasting blood samples were analysed using enzymatic techniques on a Hitachi 747 analyser to measure cholesterol and triglycerides. HDL-C was determined by enzymatic assay after polyethylene glycol precipitation, and LDL-C was estimated using the Friedewald formula [S1]. Blood pressure measurements included systolic blood pressure (SBP; mmHg) and diastolic blood pressure (DBP; mmHg), each averaged from three readings. Glucose metabolism traits included glucose (mmol/L), insulin (mU/L), and homeostatic model assessment of insulin resistance (HOMA-IR; unitless). Medical history and lifestyle variables were obtained from self-reported questionnaires. Diabetes status was defined using self-reported diabetes information and reviewed for consistency with available glucose metabolism measures, including HOMA-IR. Lipid-lowering medication use was based on self-reported questionnaire data. Smoking status was defined as ever smoking, based on self-reported smoking of at least seven cigarettes per week, one cigar per week, or one ounce of tobacco per month for as long as one year.

Candidate covariates for the final adjusted models were selected based on cardiovascular relevance, availability in the 1994-95 BHS dataset, consistency with established cardiovascular risk-prediction approaches, and model-fit assessment in the broader BHS female dataset before exclusion of women with prevalent cardiovascular disease (CVD). Candidate predictors were assessed within broad phenotype categories, including anthropometrics, blood pressure, glucose metabolism, and lipids. Age and age^2^ were included in all candidate models. Model fit was compared using Akaike information criterion (AIC), with lower AIC indicating better model fit.

Within the anthropometric category, waist circumference provided the best-fitting model after adjustment for age and age^2^ (AIC = 1764.55; odds ratio [OR] 1.02, 95% confidence interval [CI] 1.01-1.03; p = 0.0004). Within the glucose metabolism category, diabetes status had the lowest AIC (AIC = 1770.35; OR 5.09, 95% CI 1.44-32.36; p = 0.031). Within the lipid category, TC/HDL-C ratio had the lowest AIC (AIC = 1773.13; OR 1.11, 95% CI 1.00-1.23; p = 0.042). These variables were therefore retained as the adiposity, glucose metabolism, and lipid measures in the final adjusted models.

For blood pressure, DBP had the lowest AIC in the age- and age^2^-adjusted candidate model (AIC = 1774.26; OR 1.01, 95% CI 1.00-1.02; p = 0.080). However, the corresponding SBP model had very similar fit, with a small difference in AIC compared with DBP (AIC = 1775.32; OR 1.01, 95% CI 1.00-1.01; p = 0.156). Because this difference in model fit was small, SBP was selected as the final blood pressure covariate to align with established CVD risk-prediction approaches, particularly the modified PREDICT New Zealand CVD risk equation [S2]. The PREDICT-1° equation includes age, smoking status, diabetes status, SBP, TC/HDL-C ratio, and preventive pharmacotherapy, including lipid-lowering medication use, as predictors, which overlapped with the main cardiometabolic covariates selected for the present study [S2]. The broader PREDICT equation also includes sex, ethnicity, socioeconomic deprivation, family history of CVD, history of atrial fibrillation, and other preventive medications; however, these were not the focus of the phenotype category selection undertaken in the present analysis [S2].

The final covariate set included age, age^2^, SBP, waist circumference, TC/HDL-C ratio, diabetes status, lipid-lowering medication use, smoking status, and the first ten genetic principal components. These covariates were used consistently in the fully adjusted Fine-Gray competing-risk models.

**Supplementary Methods S2. Genotype quality control and imputation**

Genotyping was performed using the Illumina Human 610K Quad-Bead Chip at the Centre National de Genotypage, Paris, and the Illumina 660W Quad-Array Bead Chip at PathWest Laboratory Medicine, Western Australia. Genotype quality control (QC) was conducted before imputation using PLINK v1.9/2.0 [S3,S4].

Sample-level QC excluded individuals with genotype missingness greater than 3%, sex discrepancies, heterozygosity outliers, ancestry outliers, and duplicate samples where identified. Variant-level QC excluded single nucleotide polymorphisms (SNPs) with missingness greater than 3%, duplicate variants, variants failing minor allele frequency filtering, variants deviating from Hardy-Weinberg equilibrium (p < 5.0 x 10^-4^), ambiguous genotypes, and variants with mismatches to the Haplotype Reference Consortium (HRC) reference panel [S5-S8].

Genomic coordinates were lifted from GRCh36/hg18 to GRCh38/hg38 using the University of California Santa Cruz (UCSC) LiftOver tool [S5]. Pre-imputation processing was performed using the Will Rayner checking tools [S6]. This included allele matching, strand-orientation checks, comparison of allele frequencies with reference data, removal of indels and duplicate variants, and exclusion of variants with unresolved allele mismatches or strand inconsistencies. Database of Single Nucleotide Polymorphisms (dbSNP) resources were used to support variant annotation and allele-frequency checking [S7].

Genotype imputation was performed using the Trans-Omics for Precision Medicine (TOPMed) Imputation Server [S9,S11]. Eagle2 was used for phasing [S10], and Minimac4 was used for imputation [S11]. After imputation, post-imputation QC retained variants with imputation quality R^2^ > 0.5 and minor allele frequency ≥ 0.001. Multiallelic variants and indels were excluded. The post-imputation dataset after QC was used for downstream polygenic risk score calculation.

Population structure was assessed using principal component analysis (PCA) in PLINK 2.0 [S4]. The first ten genetic principal components were retained and included as covariates in the fully adjusted models to account for residual population structure.

**Supplementary Methods S3. Genetic ancestry assessment and polygenic risk score workflow**

Selection of hypertensive disorders of pregnancy (HDP)-related polygenic risk scores (PRS) was restricted to externally derived scores with available scoring weights and sufficient compatibility with the Busselton Health Study (BHS) imputed genotype data. The selected scores included two preeclampsia (PE) PRS, PGS003586 and PGS004593, one gestational hypertension (GH) PRS, PGS003587, and one broad HDP PRS that was not available in the Polygenic Score (PGS) Catalog. The BHS 1994-95 post-imputation quality-controlled genotype dataset included 12,911,694 genetic variants. Variant overlap between each score and the BHS post-imputation dataset was assessed before downstream analysis. Scores were retained if at least 75% of variants in the original scoring file were available after harmonisation with the BHS imputed genotype data.

Score harmonisation included matching variant identifiers, checking genome build compatibility, aligning effect alleles, and removing variants that could not be reliably matched to the BHS genotype data. For the three PGS Catalog scores, harmonisation and score calculation were performed using the PGS Catalog Calculator, pgsc_calc v2.2.0 [S12,S13]. The broad HDP PRS was processed using the same harmonised genotype data and the published score weights from its source file.

Genetic ancestry was assessed within the pgsc_calc pipeline using FRAPOSA (Fast and Robust Ancestry Prediction using Online Singular Value Decomposition and Shrinkage Adjustment) [S13,S14]. BHS participants were compared with Human Genome Diversity Project (HGDP) and 1000 Genomes Project reference populations, including African, American, Central/South Asian, East Asian, European, and Middle Eastern reference groups [S15,S16]. The workflow generated principal component analysis plots and ancestry similarity summaries to assess genetic similarity between BHS participants and reference populations. These outputs were used to support interpretation of PRS transferability, particularly because the included HDP-related PRS were predominantly derived from European-ancestry source populations.

The pgsc_calc workflow produced multiple PRS output formats, including Z-score-normalised outputs [S13]. However, for the primary statistical analyses, raw sum scores were retained and then standardised within the final BHS analysis cohort, allowing all PRS effect estimates to be interpreted consistently per one standard deviation increase in score.

Analyses were executed using Nextflow on the Pawsey Supercomputing system to support reproducible and scalable workflow execution [S17]. The workflow was run within a Singularity container [S18]. Singularity is a containerisation platform commonly used on high-performance computing systems; it packages the required software, dependencies, and runtime environment into a portable container, allowing the same workflow to be run consistently across computing environments without requiring administrative privileges. Supporting tools included PLINK 2.0 for genotype processing and pygscatalog for PRS retrieval and integration [S4,S13].

Further details on the source and target phenotype, source publication, development population, original and analysis genome builds, number of variants in the original score file, variant matching with the BHS genotype data, and calculation method for each PRS are provided in Supplementary Table S2.

| **Supplementary Table S2. Polygenic risk scores included in the analysis** | | | | | | | | | |
| --- | --- | --- | --- | --- | --- | --- | --- | --- | --- |
| **Score ID / source file** | **Target phenotype** | **Source publication / resource** | **Development/**  **source population** | **Source PRS development method** | **Official original genome build** | **Build used in BHS workflow** | **Original variants in score file** | **Matched variants in BHS, n (%)** | **Unmatched variants in BHS** |
| PGS003586 | Preeclampsia | Honigberg et al., 2023 [S19] | Variant associations from multi-ancestry GWAS; score development/training in European participants | PRS-CS [S23] | GRCh37 | Harmonised GRCh38 PGS Catalog score file | 1,087,033 | 1,083,754 (99.70%) | 3279 |
| PGS003587 | Gestational hypertension | Honigberg et al., 2023 [S19] | Variant associations from multi-ancestry GWAS; score development/training in European participants | PRS-CS | GRCh37 | Harmonised GRCh38 PGS Catalog score file | 1,087,916 | 1,085,264 (99.76%) | 2652 |
| PGS004593 | Preeclampsia | Nurkkala et al., 2023 [S20] | Multi-ancestry source GWAS including European participants; evaluated in European/Finnish samples | PRS-CS | GRCh38 | Harmonised GRCh38 PGS Catalog score file | 1,102,059 | 1,099,489 (99.77%) | 2570 |
| Broad HDP PRS:  O15_HYPTENSPREG_FinnGen_r8_PRScs_auto | Hypertensive disorders of pregnancy | Kim Lab GitHub repository [S21]; Jung et al., 2025 [S22] | FinnGen R8-derived HDP PRS; PRS-CS using 1000 Genomes Phase 3 European LD reference | PRS-CS | GRCh37/hg19 source score file | Lifted/harmonised to GRCh38 for BHS analysis | 1,102,676 | 1,100,412 (99.79%) | 2264 |

**Note: BHS**, Busselton Health Study; **GWAS**, genome-wide association study; **HDP**, hypertensive disorders of pregnancy; **LD**, linkage disequilibrium; **PGS**, polygenic score; **PRS**, polygenic risk score; **PRS-CS**, polygenic risk score continuous shrinkage method. Source PRS development method refers to the method used in the original publication or resource to generate PRS weights. Official original genome build refers to the genome build reported in the PGS Catalog or source score file. The three PGS Catalog scores were calculated using harmonised GRCh38 score files in the BHS workflow, whereas the broad HDP PRS was supplied as a GRCh37/hg19 source score file and lifted/harmonised to GRCh38 for BHS analysis. Variant matching was assessed against the BHS 1994-95 post-imputation genotype dataset after harmonisation. Raw sum scores were retained and standardised within the final analysis cohort for analysis.

**Supplementary References**

[S1] W.T. Friedewald, R.I. Levy, D.S. Fredrickson, Estimation of the concentration of low-density lipoprotein cholesterol in plasma, without use of the preparative ultracentrifuge, Clin. Chem. 18 (1972) 499-502. <https://doi.org/10.1093/clinchem/18.6.499>.

[S2] R. Pylypchuk, S. Wells, A. Kerr, K. Poppe, T. Riddell, M. Harwood, et al., Cardiovascular disease risk prediction equations in 400 000 primary care patients in New Zealand: a derivation and validation study, Lancet 391 (2018) 1897-1907. <https://doi.org/10.1016/S0140-6736(18)30664-0>.

[S3] S. Purcell, B. Neale, K. Todd-Brown, L. Thomas, M.A.R. Ferreira, D. Bender, et al., PLINK: a tool set for whole-genome association and population-based linkage analyses, Am. J. Hum. Genet. 81 (2007) 559-575. <https://doi.org/10.1086/519795>.

[S4] C.C. Chang, C.C. Chow, L.C.A.M. Tellier, S. Vattikuti, S.M. Purcell, J.J. Lee, Second-generation PLINK: rising to the challenge of larger and richer datasets, Gigascience 4 (2015) 7. <https://doi.org/10.1186/s13742-015-0047-8>.

[S5] A.S. Hinrichs, D. Karolchik, R. Baertsch, G.P. Barber, G. Bejerano, H. Clawson, et al., The UCSC Genome Browser Database: update 2006, Nucleic Acids Res. 34 (2006) D590-D598. <https://doi.org/10.1093/nar/gkj144>.

[S6] N.W. Rayner, HRC or 1000G Imputation Preparation and Checking, Will Rayner pre-imputation checking tools. <https://www.chg.ox.ac.uk/~wrayner/tools/> (accessed 7 July 2026).

[S7] S.T. Sherry, M.H. Ward, M. Kholodov, J. Baker, L. Phan, E.M. Smigielski, et al., dbSNP: the NCBI database of genetic variation, Nucleic Acids Res. 29 (2001) 308-311. <https://doi.org/10.1093/nar/29.1.308>.

[S8] S. McCarthy, S. Das, W. Kretzschmar, O. Delaneau, A.R. Wood, A. Teumer, et al., A reference panel of 64,976 haplotypes for genotype imputation, Nat. Genet. 48 (2016) 1279-1283. <https://doi.org/10.1038/ng.3643>.

[S9] D. Taliun, D.N. Harris, M.D. Kessler, J. Carlson, Z.A. Szpiech, R. Torres, et al., Sequencing of 53,831 diverse genomes from the NHLBI TOPMed Program, Nature 590 (2021) 290-299. <https://doi.org/10.1038/s41586-021-03205-y>.

[S10] P.R. Loh, P. Danecek, P.F. Palamara, C. Fuchsberger, Y.A. Reshef, H.K. Finucane, et al., Reference-based phasing using the Haplotype Reference Consortium panel, Nat. Genet. 48 (2016) 1443-1448. <https://doi.org/10.1038/ng.3679>.

[S11] S. Das, L. Forer, S. Schönherr, C. Sidore, A.E. Locke, A. Kwong, et al., Next-generation genotype imputation service and methods, Nat. Genet. 48 (2016) 1284-1287. <https://doi.org/10.1038/ng.3656>.

[S12] S.A. Lambert, L. Gil, S. Jupp, S.C. Ritchie, Y. Xu, A. Buniello, et al., The Polygenic Score Catalog as an open database for reproducibility and systematic evaluation, Nat. Genet. 53 (2021) 420-425. <https://doi.org/10.1038/s41588-021-00783-5>.

[S13] S.A. Lambert, B. Wingfield, J.T. Gibson, L. Gil, S. Ramachandran, F. Yvon, et al., Enhancing the Polygenic Score Catalog with tools for score calculation and ancestry normalization, Nat. Genet. 56 (2024) 1989-1994. <https://doi.org/10.1038/s41588-024-01937-x>.

[S14] D. Zhang, R. Dey, S. Lee, Fast and robust ancestry prediction using principal component analysis, Bioinformatics 36 (2020) 3439-3446. <https://doi.org/10.1093/bioinformatics/btaa152>.

[S15] L.L. Cavalli-Sforza, The Human Genome Diversity Project: past, present and future, Nat. Rev. Genet. 6 (2005) 333-340. <https://doi.org/10.1038/nrg1596>.

[S16] 1000 Genomes Project Consortium, A global reference for human genetic variation, Nature 526 (2015) 68-74. <https://doi.org/10.1038/nature15393>.

[S17] P. Di Tommaso, M. Chatzou, E.W. Floden, P.P. Barja, E. Palumbo, C. Notredame, Nextflow enables reproducible computational workflows, Nat. Biotechnol. 35 (2017) 316-319. <https://doi.org/10.1038/nbt.3820>.

[S18] G.M. Kurtzer, V. Sochat, M.W. Bauer, Singularity: scientific containers for mobility of compute, PLoS One 12 (2017) e0177459. <https://doi.org/10.1371/journal.pone.0177459>.

[S19] M.C. Honigberg, B. Truong, R.R. Khan, B. Xiao, L. Bhatta, H.M.T. Vy, et al., Polygenic prediction of preeclampsia and gestational hypertension, Nat. Med. 29 (2023) 1540-1549. <https://doi.org/10.1038/s41591-023-02374-9>.

[S20] J. Nurkkala, A. Kauko, FinnGen, H. Laivuori, T. Saarela, J.S. Tyrmi, et al., Associations of polygenic risk scores for preeclampsia and blood pressure with hypertensive disorders of pregnancy, J. Hypertens. 41 (2023) 380-387. <https://doi.org/10.1097/HJH.0000000000003336>.

[S21] Dokyoon Kim Lab, PRS model for hypertensive disorders of pregnancy: hdp-prs-finngen-r8, Zenodo. <https://doi.org/10.5281/zenodo.14211027> (accessed 29 July 2026).

[S22] S.H. Jung, H. Kim, Y.M. Jung, M. Shivakumar, B. Xiao, J. Kim, et al., Healthy lifestyle reduces cardiovascular risk in women with genetic predisposition to hypertensive disorders of pregnancy, Nat. Commun. 16 (2025) 1463. <https://doi.org/10.1038/s41467-025-56107-2>.

[S23] T. Ge, C.Y. Chen, Y. Ni, Y.A. Feng, J.W. Smoller, Polygenic prediction via Bayesian regression and continuous shrinkage priors, Nat. Commun. 10 (2019) 1776. <https://doi.org/10.1038/s41467-019-09718-5>.
